# Identifying novel high-impact rare disease-causing mutations, genes and pathways in exomes of Ashkenazi Jewish inflammatory bowel disease patients

**DOI:** 10.1101/2020.07.01.20143750

**Authors:** Yiming Wu, Kyle Gettler, Mamta Giri, Dalin Li, Cigdem Sevim Bayrak, Aayushee Jain, Patrick Maffucci, Ksenija Sabic, Tielman Van Vleck, Girish Nadkarni, Lee A. Denson, Harry Ostrer, Subra Kugathasan, Peter D. Stenson, David N. Cooper, L. Philip Schumm, Scott Snapper, Mark J. Daly, Talin Haritunians, Richard H. Duerr, Mark S. Silverberg, John D. Rioux, Steven R. Brant, Dermot McGovern, Judy H. Cho, Yuval Itan

## Abstract

Inflammatory bowel disease (IBD) is a group of chronic diseases, affecting different parts of the gastrointestinal tract, that mainly comprises Crohn’s Disease (CD) and Ulcerative Colitis (UC). Most IBD genomic research to date has involved genome-wide association studies (GWAS) of common genetic variants, mostly in Europeans, resulting in the identification of over 200 risk loci. The incidence of IBD in Ashkenazi Jews (AJ) is particularly high compared to other population groups and rare protein-coding variants are significantly enriched in AJ. These variants are expected to have a larger phenotypic effect and are hypothesized to complement the missing heritability that cannot be fully addressed by GWAS in IBD. Therefore, we genetically identified 4,974 AJs IBD cases and controls from whole exome sequencing (WES) data from the NIDDK IBD Genetics Consortium (IBDGC). We selected credible rare variants with high predicted impact, aggregated them into genes, and performed gene burden and pathway enrichment analyses to identify 7 novel plausible IBD-causing genes:*NCF1, CES1, ICAM1, INPP5D, ABCB1, IL33* and *TLR4*. We further perform bulk and single-cell RNA sequencing, demonstrating the likely relatedness of the novel genes to IBD. Importantly, we demonstrate that the rare and high impact genetic architecture of AJ adult IBD displays a significant overlap with very early onset IBD (VEOIBD) genetics. At the variant level, we performed Phenome-wide association studies (PheWAS) in the UK Biobank to replicate risk sites in IBD and reveal shared risk sites with other diseases. Finally, we showed that a polygenic risk score (PRS) has high power to differentiate AJ IBD cases from controls when using rare and high impact variants.

## MAIN (Introduction, Results and Discussion)

Inflammatory bowel disease (IBD) is a group of chronic diseases where sections of the gastrointestinal tract are inflamed due to an aberrant immune response to intestinal bacteria and microbiota in genetically susceptible individuals. IBD comprises mainly Crohn’s Disease (CD) and Ulcerative Colitis (UC). Genome-wide association studies (GWAS) have identified more than 200 IBD risk loci to date, mostly in Europeans^1-4^. The Ashkenazi Jewish (AJ) population has a high IBD susceptibility, with a 2- to 4-fold increased risk of developing IBD due to an AJ founder effect and long-term genetic isolation^5-7^. A recent study indicated that 34% of rare protein coding variants present in the AJ population are significantly enriched in comparison to other reference populations^8^. Therefore rare and high impact genetic variants in AJ may address and complement the missing heritability of current IBD GWAS studies^9^.

In this study, we genetically identified 4,974 QC-passed AJs IBD cases and controls from whole exome sequencing (WES) of the NIDDK IBD Genetics Consortium (IBDGC). We employed several cutting-edge approaches to select highly credible rare variants which are predicted to have high phenotypic impact, then performed a SNP-set Kernel Association Test (SKAT) on gene-level aggregations of these high impact rare variants. In addition, we performed meta and pathway enrichment analyses to identify novel plausible IBD-causing candidate genes which we further validated by bulk RNA sequencing (RNA-seq) and single-cell RNA sequencing (scRNA-seq) analyses. At the variant level, we conducted PheWAS analyses to replicate risk sites on IBD and to discover shared risk sites with other diseases. Finally, we tested the polygenic risk score (PRS) classification performance of filtered high impact variants in IBD cases and controls by using a Random Forest machine learning algorithm.

### Genetically identifying Ashkenazi Jewish samples

To perform a high-quality gene burden case-control analysis, it is important to focus on homogenous and genetically matched cases and controls. We first genetically identified samples of AJ ancestry across all 9,076 IBDGC WES samples. We performed population structure admixture analysis to estimate the AJ fraction of each sample. We then used principal component analysis (PCA) to validate the genetically identified AJs by comparing them to the AJ reference panel^6^. We filtered a set of LD-pruned independent sites to perform a fastSTRUCTURE admixture analysis by comparing individual samples with 36 known AJ reference samples^10^. The lowest AJ fraction (0.645) in the AJ reference panel was used as the threshold, above which a WES sample was determined to be genetically AJ and retained for further analyses (Supplementary Fig. 2, Methods). 1,744 AJ WES samples were identified from IBDGC dataset 1, forming a distinct cluster overlapping the AJ reference panel, and quite distinct from the European samples in the PCA analysis (Fig. 1b). We applied the same AJ identification process to the IBDGC dataset 2 to genetically identify 3,338 AJ WES samples. After a quality control (QC) process (Methods), we collectively obtained 4,974 AJ samples, comprising 1,905 IBD AJ cases (1,258 CD, 496 UC, and 151 IBD) and 3,069 AJ controls.

**Figure 1.**
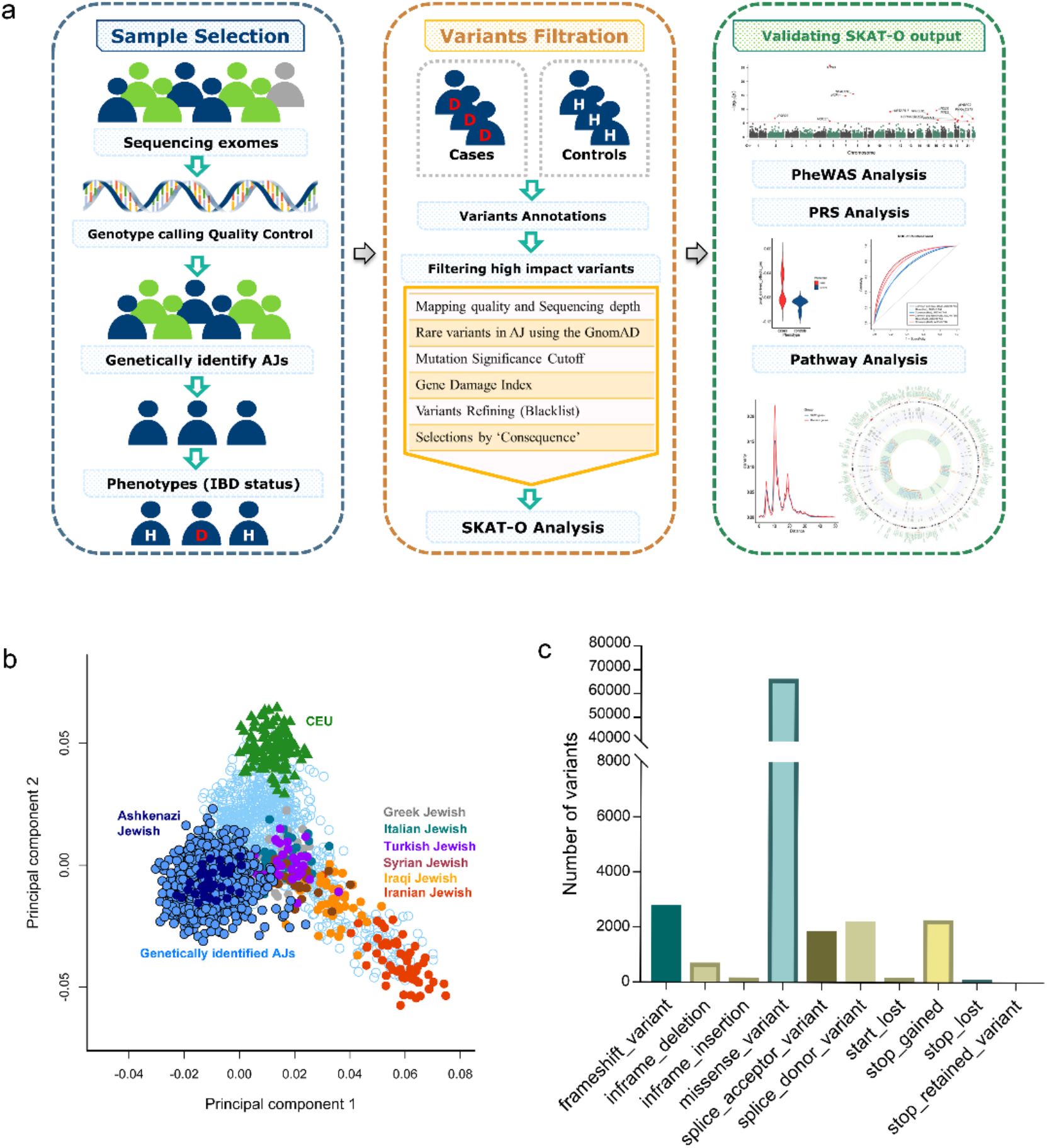
**a**. Flowchart of present work. Firstly, AJ samples from all WES participants that passed quality control were genetically identified. Then high impact rare variants from exomes and flanking regions were filtered using cutting-edge mutation filtering approaches and high impact rare variants were aggregated into gene sets to perform SKAT-O analysis on IBD cases and controls of AJs. Finally, IBD associations were validated and prioritized at pathway level, gene level and variant level using multiple methods. **b**. Genetically identified Ashkenazi Jewish samples are displayed on a PCA plot compared to the Jewish and European reference panels. The genetically identified AJs display an independent cluster, which overlapped the AJ reference panel and was distinct from the European cluster. **c**. Distributions of filtered high impact rare variants by molecular function.

### IBD candidate genes identification from high impact rare variants

To identify plausible IBD-causing candidate genes, we first performed a SNP-set Kernel Association Test (SKAT)^11^ on gene-level aggregations of filtered high impact variants, obtained by several cutting-edge approaches to select highly credible deleterious variants (Methods and Fig. 1a). We performed AJ IBD case-control SKAT-O analysis on 14,589 genes harboring 96,309 high impact rare variants (Fig. 1c), then performed SKAT-O analyses of IBD, CD and UC cases versus unaffected controls. We identified 15 genes displaying genome-wide significance (Bonferroni corrected *P* = 3.42×10^−6^ (=0.05/14,589)), that included the well-described CD gene *NOD2* (Fig. 2a, Supplementary Table 1). *NOD2* had higher significance in the CD-specific SKAT-O analysis (*P=*9.87×10^−16^, Supplementary Fig. 3. Supplementary Table 2) but was insignificant in the UC-specific analysis (*P=*1) (Supplementary Fig. 4, Supplementary Table 3) as expected, since *NOD2*^12^ is not known to cause UC. To examine the contribution of variants within the significant genes, we performed SKAT-O single variant tests on AJ IBD cases versus AJ unaffected controls (Supplementary Fig. 5) where most significant genes harbor one or more variants passing the genome-wide association threshold (*P* = 5×10^−8^). In addition, we performed a logistic regression association analysis on all high impact variants in AJ IBD cases and AJ controls, see Supplementary Table 4 for variants with *P* value < 0.05 and their host genes. At the gene level, the top ranking significant genes (with *P* < 0.01) were selected for further pathway analyses.

**Figure 2.**
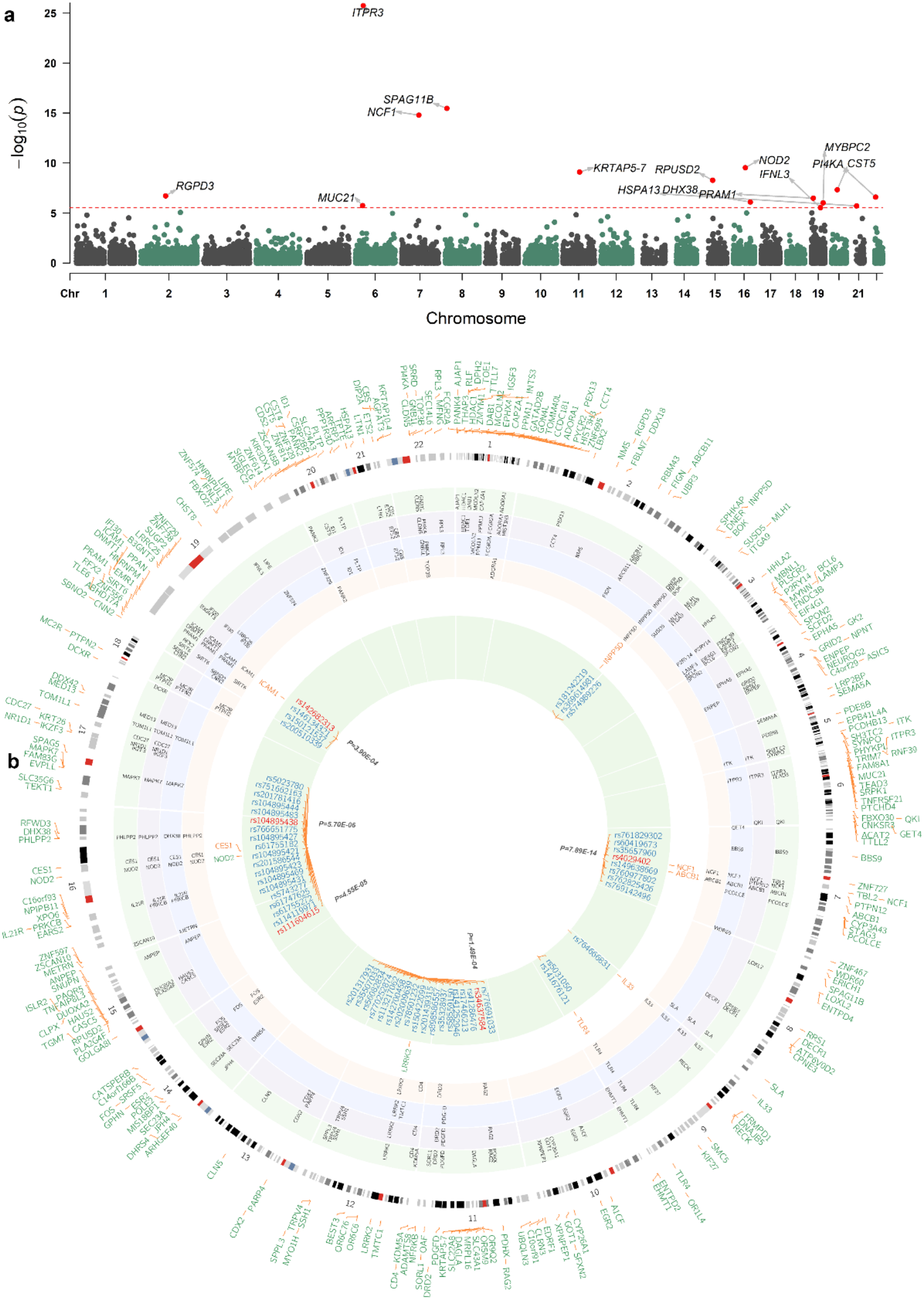
**a**. SKAT-O analysis on 1,905 AJ IBD cases and 3,069 AJ controls. The red dashed line indicates the Bonferroni adjusted *P* values of genome-wide significance. **b**. A circos plot summarizes the process of identifying IBD-associated genes and variants. The outer layer includes all 268 SKAT-O derived IBD candidate genes with *P* < 0.01. The intermediate four layers represent the top genes identified by four different pathway enrichment and biological relatedness analyses (green: ToppGene; purple: Human Gene Connectome; blue: GIANT; yellow: Ingenuity Pathway Analysis). The 9 genes listed between intermediate layers and inner green layer are significant genes commonly identified by all four pathway approaches (two well-known IBD genes in green and 7 novel genes in orange). The inner layer displays all 55 high-impact rare variants in the 9 orange genes. Of the 55 variants, 5 variants (highlighted in red inside the inner green layer) are associated with IBD.

### Pathway enrichment and biological relatedness analyses

To validate the significant IBD-associated genes from the SKAT-O case-control analyses, we performed pathway enrichment and biological relatedness analyses to identify and prioritize plausible IBD-causing candidate genes. Genes with *P*-value < 0.01 in the AJ IBD SKAT-O test were used as candidates (268 genes, of which 5 are known IBD associated genes: *NOD2, FCGR2A, PRKCB, LRRK*2, and *FOS*), from which a subset of genes was identified by 4 pathway enrichment and biological relatedness methods: Ingenuity Pathway Analysis (IPA), ToppGene, Genome-Scale Integrated Analysis of Networks in Tissues (GIANT), and Human Gene Connectome (HGC, Methods). First, we used IPA to identify candidate gene-related pathways, diseases and biological functions^13^. The ‘Gastrointestinal Disease’ term was ranked 3rd under the ‘Top Disease and Biological Functions’ category (the top two terms being ‘Cancer’ and ‘Organismal Injury and Abnormalities’, where it has been shown that IBD patients have a higher risk for developing several types of cancer^14^). Of the 56 sub-functions of ‘Gastrointestinal Disease’, we collated 22 genes from three function modules: inflammation of gastrointestinal tract (*P* = 1.98×10^−5^), inflammation of small intestine (*P* = 3.41×10^−4^) and colitis (*P* = 4.74×10^−4^), which are implicated in IBD (Methods, Supplementary Table 5). We then employed ToppGene to rank candidate genes according to their relatedness to known IBD genes based on functional annotation and protein interaction network^15^. The candidate genes were sorted by an overall *P*-value generated by integrating functional annotation and protein interactions network. 113 genes with *P* < 0.05 were selected as top candidates (Methods, Supplementary Table 5). We next used the module detection tool GIANT to identify IBD-related genes from our candidate genes^16^. Using GIANT’s default setting (global tissue), we identified five functional modules, of which one was highly correlated with immunological function, from which we extracted its 60 genes (Methods, Supplementary Table 5).

Finally, we used HGC to estimate whether the IBD candidate genes are significantly related to known IBD-associated genes (Methods)^17^ in comparison to randomly selected human genes. For each candidate gene, we calculated its average HGC biological proximity to 157 known IBD-causing gene and compared to randomly sampled 268 genes in 10,000 re-sampling iterations, obtaining a *P*-value of 0.031, (Methods, Fig. 3b.c, Supplementary Fig. 6), demonstrating a significant functional association of the candidate genes to IBD. We then tested the biological association to the list of known very early onset inflammatory bowel disease (VEO-IBD)-causing genes, resulting in a *P* value of 0.023 compared to random gene sets in 10,000 resampling iterations. These analyses indicate that, under the model of rare high impact mutations, adult IBD genetics resembles that of IBD of young children. In addition, we applied functional genomic alignment (FGA) to cluster all candidate genes with known IBD genes by their biological distance (Fig. 3a). The candidate genes were evenly intermixed with the known IBD genes, indicating that the candidate IBD genes are likely to be associated with the IBD phenotype. Finally, we obtained 73 genes with an average HGC distance lower than 11.13 (a cutoff based on the average distance between known IBD-associated genes) for further analyses (Methods, Supplementary Table 5). We identified a final list of nine genes (*NCF1, CES1, ICAM1, INPP5D, ABCB1, IL33, TLR4, NOD2*, and *LRRK2*) that occurred in the top ranking results from all 4 pathway enrichment and biological distance approaches. Of these genes, 4 have not been reported as having pathogenic mutations in the Human Gene Mutation Database (HGMD) Professional version (*ICAM1, INPP5D, IL33* and *TLR4*)^18^, and 3 genes are reported as having pathogenic mutations in non-IBD diseases (*NCF1* in chronic granulomatous disease, *CES1* in carboxylesterase 1deficiency, and *ABCB1* in Parkinson disease) in HGMD. The remaining two genes, *NOD2* and *LRRK2*, are well known IBD genes (Table 1), whereas the other 7 are novel genes yet to be formally implicated in IBD studies (Fig. 2b). In comparison with our gene prioritization results, which are generated by integrating results from more pathway and functional enrichment analyses (Methods), six genes remained in the top 10% (Top 26 of 268 genes) of the prioritized gene list: *NOD2, TLR4, LRRK2, ICAM1, INPP5D*, and *NCF1* in descending order regarding predicted functional relatedness to IBD (Supplementary Table 6, Supplementary Fig. 7). All 6 genes display strong biological relevance to IBD: *TLR4* plays a key role as the hub of the immune response to microbes in the gut in IBD pathogenesis^19^. *ICAM1* is involved in adhesive interaction mediation between the lymphocytes and endothelial cells, and has been recognized as a therapeutic target in IBD^20,21^. To assess whether the *ICAM1* top variant (rs142682313, OR=0.391, *P*=3.90×10^−04^) is conditionally independent of IBD associated sites in *TYK2*^22^, we performed joint and conditional analyses using GCTA-COJO with *ICAM1* lead SNP and three IBD-associated sites in *TYK2*, both of which suggested it has independent protective effects against IBD (Supplementary Table 7, Supplementary Table 8). *INPP5D* encodes SHIP proteins, whose expression level is significantly associated with IBD^23,24^. *INPP5D* resides in close proximity to another IBD gene, *ATG16L1*^25^. We therefore conducted a linkage disequilibrium analysis on the most significant variant in *INPP5D*: rs574989226 (*P*=0.011) and demonstrated that there were no strong LD pairs identified between *ATG16L1* and *INPP5D* (Supplementary Table 9). The *NCF1* protein is an essential component of the phagocytic Nicotinamide Adenine Dinucleotide Phosphate (NADPH) oxidase complex type 2 (the NOX2 complex), which produces anti-inflammatory reactive oxygen species (ROS) and prevents autoimmune responses^26^. Variants in the NOX2 NADPH oxidase complex have been found to confer susceptibility to VEO-IBD^27^. *IL33* has long been considered to play an important role in intestinal immunity. *IL33* and its membrane receptor *ST2* act as a critical regulator of inflammation^28,29^. *ABCB1* (also known as *MDR1*) was considered as a VEO-IBD related gene^30^. However, while no strong evidence yet supports its role in IBD pathogenesis, experiments with MDR1a knockout mice have indicated that *MDR1* deficiency can cause colitis^31,32^. In addition, few variants (Ile1145Ile, rs1045642; Ala893Ser/Thr, rs2032582) have been reported as having modest effects in previous case-control analyses^33,34^. No previous studies have associated *CES1* with IBD. *CES1* is an enzyme activated in the metabolic system, being a major liver enzyme which functions in liver drug clearance, including the drug Filgotinib which is utilized in CD treatment^35^.

**Table 1.**
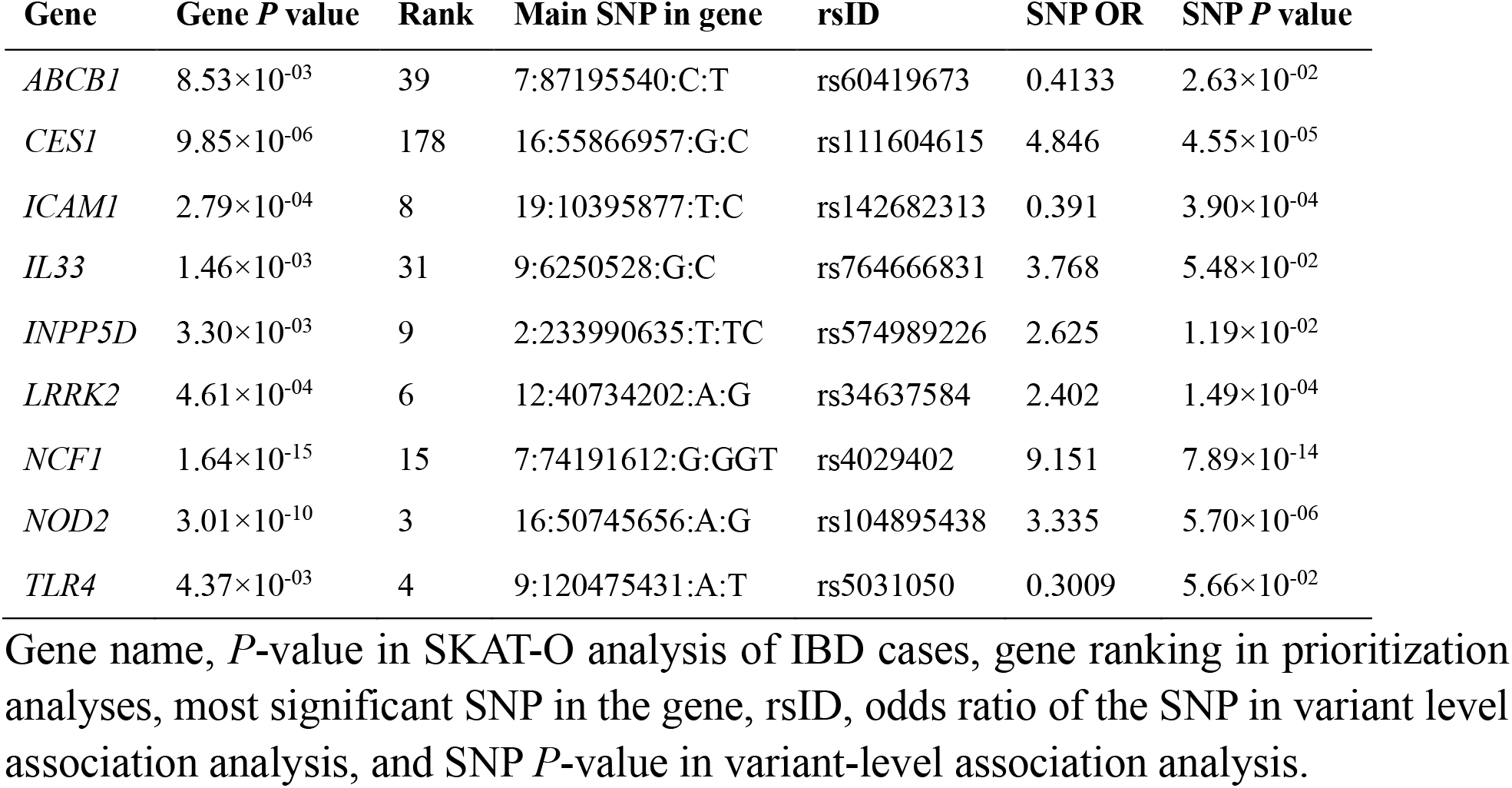
Association statistics for 9 IBD genes from pathway analyses and their lead SNPs.

| Gene | Gene *P* value | Rank | Main SNP in gene | rsID | SNP OR | SNP *P* value |
| --- | --- | --- | --- | --- | --- | --- |
| *ABCB1* | $8.53 \times 10^{-03}$ | 39 | 7:87195540:C:T | rs60419673 | 0.4133 | $2.63 \times 10^{-02}$ |
| *CES1* | $9.85 \times 10^{-06}$ | 178 | 16:55866957:G:C | rs111604615 | 4.846 | $4.55 \times 10^{-05}$ |
| *ICAM1* | $2.79 \times 10^{-04}$ | 8 | 19:10395877:T:C | rs142682313 | 0.391 | $3.90 \times 10^{-04}$ |
| *IL33* | $1.46 \times 10^{-03}$ | 31 | 9:6250528:G:C | rs764666831 | 3.768 | $5.48 \times 10^{-02}$ |
| *INPP5D* | $3.30 \times 10^{-03}$ | 9 | 2:233990635:T:TC | rs574989226 | 2.625 | $1.19 \times 10^{-02}$ |
| *LRRK2* | $4.61 \times 10^{-04}$ | 6 | 12:40734202:A:G | rs34637584 | 2.402 | $1.49 \times 10^{-04}$ |
| *NCF1* | $1.64 \times 10^{-15}$ | 15 | 7:74191612:G:GGT | rs4029402 | 9.151 | $7.89 \times 10^{-14}$ |
| *NOD2* | $3.01 \times 10^{-10}$ | 3 | 16:50745656:A:G | rs104895438 | 3.335 | $5.70 \times 10^{-06}$ |
| *TLR4* | $4.37 \times 10^{-03}$ | 4 | 9:120475431:A:T | rs5031050 | 0.3009 | $5.66 \times 10^{-02}$ |

Gene name, *P*-value in SKAT-O analysis of IBD cases, gene ranking in prioritization analyses, most significant SNP in the gene, rsID, odds ratio of the SNP in variant level association analysis, and SNP *P*-value in variant-level association analysis.

**Figure 3.**
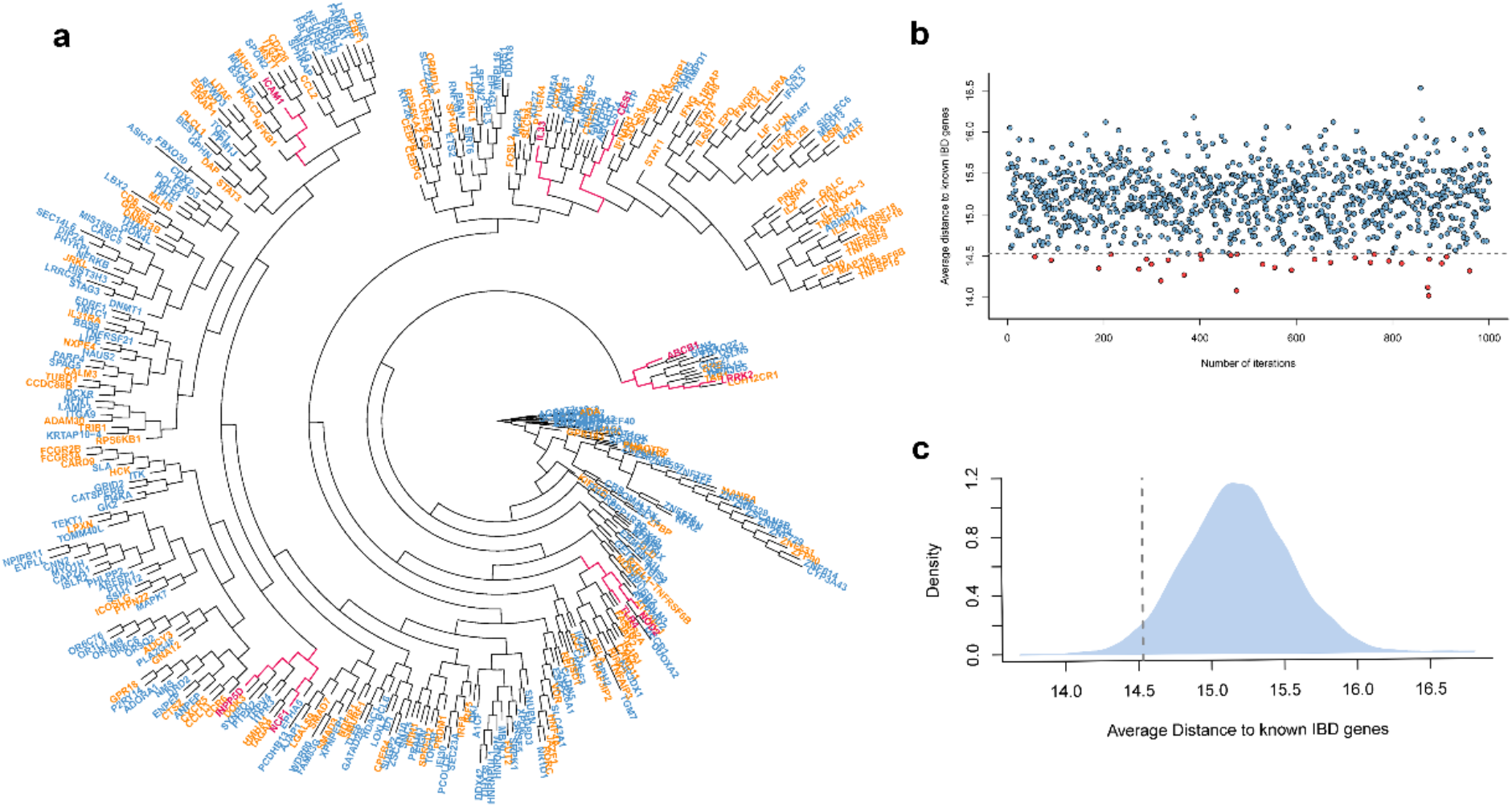
**a**. Clustering IBD candidate genes (blue) and IBD known genes (orange) according to biological relatedness by Functional Genomic Alignment function using the HGC. The 9 pathway identified genes (Fig. 2b) were emphasized in pink. The candidate genes did not form distinct clusters; rather, they are mixed with IBD known genes. **b**. A dot plot representing the average distance from a randomly selected gene set (268 genes) to known IBD genes. The grey dashed line represents the cutoff in terms of the average biological distance between IBD associated genes and IBD known genes. With genes randomly resampled 1,000 times, 31 random gene sets have lower average distances; hence, the empirical *P* value of our candidate genes being empirically associated with IBD as a group is 0.031. **c**. A density plot for all average distances in a resampling test; the percentiles at 2.5% and 97.5% are 14.50 and 15.87, respectively. The vertical dashed line denotes the cutoff in plot **b**.

To further investigate these 9 plausible IBD candidate genes, we performed single-SNP association analysis for 55 high impact variants within these genes (Supplementary Table 10). We found that each gene contained at least one high impact variant associated with IBD (*P* < 0.05). Moreover, variants in five genes passed a corrected *P*-value of 7.75×10^−4^ (rs4029402, OR=9.151, *P=*7.89×10^−14^, *NCF1*; rs104895438, OR= 3.335, *P=*5.70×10^−6^, *NOD2*; rs111604615, OR=4.846, *P=*4.55×10^−5^, *CES1*; rs34637584, OR=2.402, *P=*1.49×10^−4^, *LRRK2*; rs142682313, OR=0.391, *P=*3.9×10^−4^, *ICAM1*). rs104895438 and rs142682313 have been shown to be enriched in the AJ population and their independence has been confirmed by a previous study^8^ via conditional analyses, whereas the other variants have not been previously implicated in IBD. rs4029402 has the highest odds ratio and our finding supports its association with IBD, whilst its role and relatedness to IBD remain unclear in previous studies^36,37^. After aggregating the above 5 top ranking SNPs into a single SNP set, the mutation carrier frequency in cases was 9.9% compared to 4.6% in controls, with an odds ratio (OR) of 2.13 (*P*=7.14×10^−13^ by chi-squared test) despite a protective site that is included in the analyses.

### Gene expression RNA-seq and scRNA-seq analyses

Using RNA-seq data from IBD, CD and UC patients vs. unaffected controls^38^ we found that all 9 candidate genes are significantly over- or under-expressed in either IBD, CD or UC (Method, Fig. 4a, Supplementary Fig. 8). *INPP5D* has similar expression levels across IBD, CD and UC, where the cases display mildly lower expression than controls. *ABCB1* also has a lower expression level in cases, exhibiting lower expression in IBD and CD as compared to UC samples. We selected 268 (equal sample size as the SKAT-O significant genes) highly differentially expressed genes from IBD cases to a perform a pathway enrichment analysis in IPA. Among the results of related ‘Disease and Disorder’, it shared the ‘organismal injury and abnormalities’ with IPA results derived from the SKAT-O significant genes.

**Figure 4.**
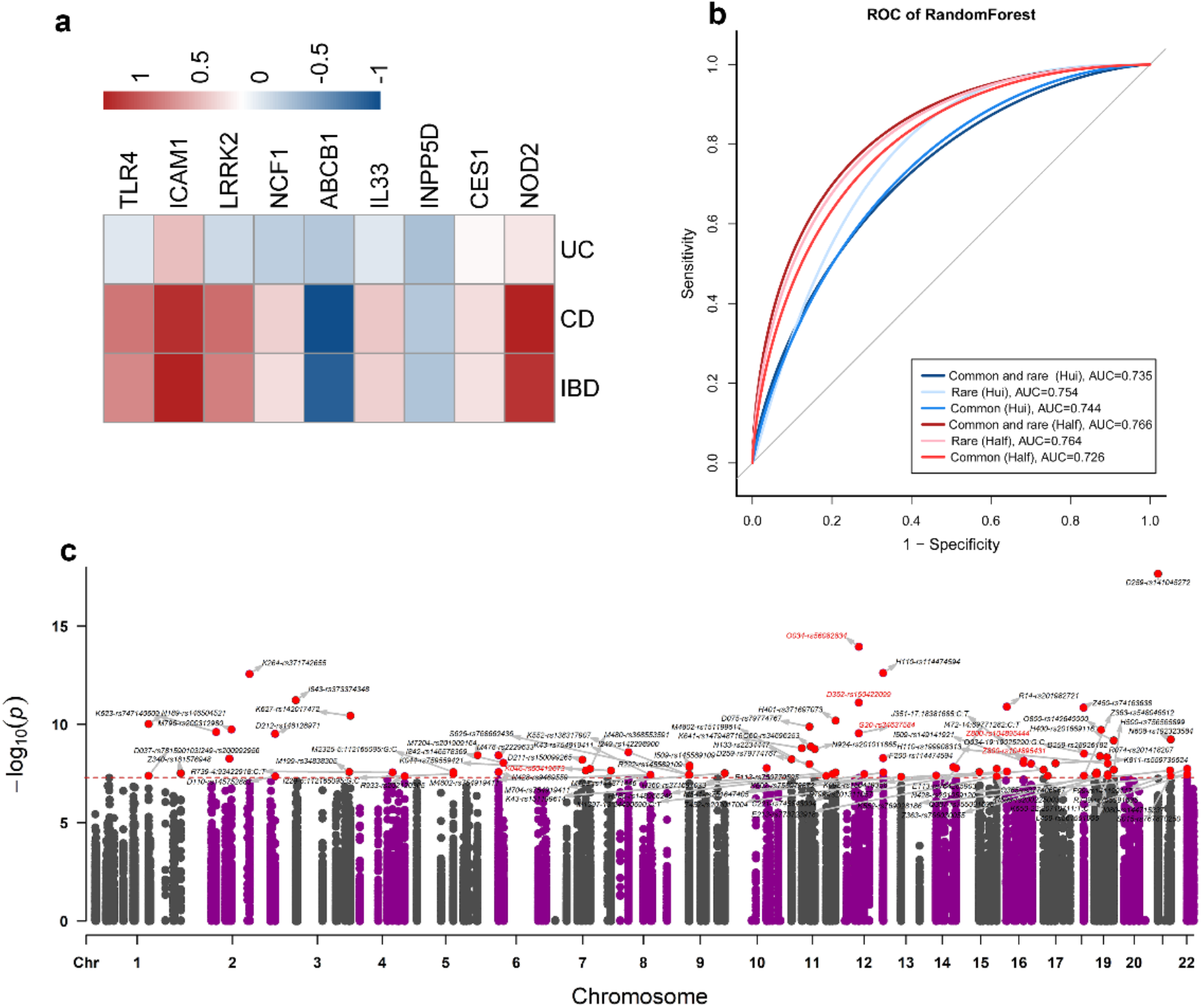
**a**. RNA-seq log-fold change of the 9 IBD associated genes identified by all four pathway analyses in CD, UC and IBD versus controls, respectively. **b**. Comparisons of prediction results on IBD individuals using polygenic risk scores derived from different variant sets (name in brackets indicating different GWAS summary statistics: Hui, Hui et al. ^45^ AJ IBD GWAS; Half, AJ IBD GWAS using half of our IBDGC samples). **c**. PheWAS analyses on high impact rare variants in SKAT-O significant IBD associated genes using 50K UK Biobank whole exome sequencing samples. ‘ICD10-Variant ID’ pairs were displayed for every significant association that passed the Bonferroni threshold (red dots). The variants pairs related to pathway identified IBD genes were displayed in red text.

Additionally, we tested gene expression data extracted from our previous scRNA-seq study of Crohn’s disease samples^39^, where the clustering of 70,226 cells from 11 paired samples (inflamed and uninflamed biopsies obtained from surgically resected ileal tissues) resulted in 36 clusters that could be annotated broadly into 16 cell-types based on the expression of specific cell type markers (Fig. 5a). We examined the expression of our 9 IBD candidate genes in the 16 different cell types in the ileum tissue. Generally, each of the 9 genes displayed over-expression in at least one cell type. *ABCB1* was mostly expressed in epithelial cells, whereas the other 8 genes showed expression in different immune system cells (Fig. 5b, Supplementary Table 11). More than half of the genes displayed over-expression in macrophages, suggesting that these genes may be involved in the immune response.

**Figure 5.**
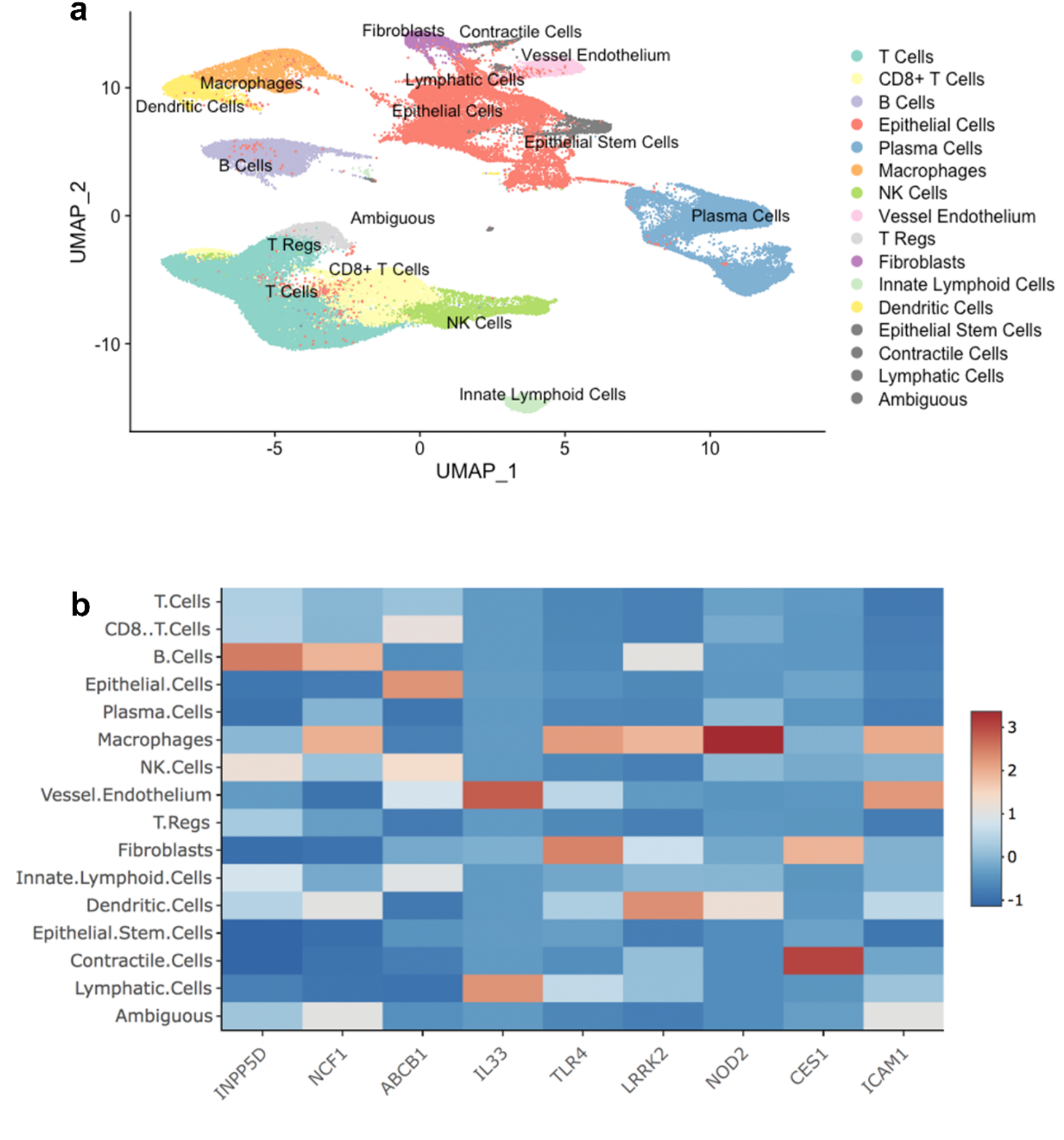
**a**. scRNA-seq of all cells from samples were grouped into 36 clusters, which can be annotated broadly into 16 cell types using Umap. **b**. scRNA-seq average log expressions of the 9 genes identified from pathway analyses across 16 cell types.

### Replication of significant genes by meta analyses

We separately tested the CD and UC associations by performing a meta-analysis of IBDGC datasets 1 and 2. Dataset 1 comprised 804 CD cases, 357 UC cases and 477 unaffected controls, whilst dataset 2 comprised 460 CD cases, 140 UC cases and 1,283 unaffected controls. Single variant associations were analyzed by raremetalworker, which provided summary statistics for the gene-level meta-analysis in RAREMETAL^40^. Of the 13,958 genes investigated, 9 genes passed the Bonferroni corrected threshold of *P* < 3.42×10^−6^ in CD case-control analysis (Supplementary Table 12). 5 of these genes were replicated from the SKAT-O CD specific analyses: *ITPR3, NOD2, CST5, MUC21* and *PRAM1*. No significant genes were identified in UC-specific meta analyses, as the unbalanced sample size between UC cases to controls dramatically decreased the power of meta-analysis^41^. *CST5* has been shown to display significant differential expression in IBD patients’ plasma levels, with higher expression in CD as compared to UC plasma^42^. *MUC21* is a primary immunodeficiency-associated gene, where variants have been shown to potentially contribute to VEO-IBD^43^.

### Phenome-wide association study analyses

The Ashkenazi Jewish population is susceptible to several complex diseases other than IBD, which prompted us to examine whether high impact variants in the IBD-associated candidate genes are shared across other traits. Therefore, we performed Phenome-wide study (PheWAS) analyses using electronic medical record (EMR) data from the UK Biobank (Methods, Supplementary Table 13). Of the 89 significant associations passing the threshold of Bonferroni adjusted *P* = 4.65×10^−8^ (Fig. 4c), we found 14 associations related to diseases of the digestive system (ICD10: K00-K95). Some of the clinical phenotypes include complications of inflammatory bowel disease, such as ‘chronic or unspecified duodenal ulcer with hemorrhage’, rs371742655, *P=*2.72×10^−13^, *ABCB11*; ‘rectal prolapse’, rs747140609, *P=*2.43×10^−10^, *LBX2*; ‘second degree hemorrhoids’, rs147948716, *P=*1.60×10^−9^, *PDHX*; ‘anal fissure; unspecified’, rs755979872, *P=*2.51×10^−8^, *ZSCAN10*; ‘angiodysplasia of colon’, rs138317907, *P=*3.64×10^−8^ and rs769028186, *P=*3.64×10^−8^ in *RECK* and *PI4KA*, respectively.

We then identified significant PheWAS associations in the genes detected by pathway analyses. Specifically, *LRRK2* has three associations: ‘Parkinson’s Disease’, rs34637584, *P=*2.80×10^−10^; ‘incomplete spontaneous abortion without complication’, rs56082834, *P=*1.12×10^−14^; ‘benign neoplasm of pituitary gland’, rs150422099, *P=*7.77×10^−12^, *NOD2* has two associations: ‘family history of malignant neoplasm of digestive organs’, rs104895431, *P=*6.78×10^−9^; rs104895444, *P=*9.24×10^−9^, *ABCB1* has one association: ‘periapical abscess with sinus’, rs60419673, *P=*9.24×10^−9^. The rs34637584 is commonly referred to as the G2019S mutation, which is a well-known Parkinson disease (G20 in ICD-10) related mutation. PheWAS results indicated that *LRRK2* can be a key gene related to the comorbidity of PD and IBD. No reports have been found to support the associations of rs5682834 to ‘incomplete spontaneous abortion without complication’ and rs150422099 to ‘benign neoplasm of pituitary gland’. Both rs104895431 and rs104895444 in *NOD2* are associated with ‘family history of malignant neoplasm of digestive organs’. The rs60419673 in *ABCB1* is associated with ‘periapical abscess with sinus’, where clinical signs of gingival inflammation have been previously found in patients with IBD^44^. For detailed PheWAS results see Supplementary Table 11.

### Polygenic risk score prediction of IBD cases using rare high impact variants

We evaluated the performance of rare and high impact variants in identifying individuals at risk for IBD using PRS with a Random Forest machine learning classification algorithm. We first used LD-pred to calculate the polygenic risk score for each individual, then employed risk scores as features to predict the IBD status of individuals with Random Forest. We have compared models based on the risk score results from six combinations of two GWAS summary statistics (a previous GWAS on AJ IBD samples from Hui et al.^45^; the other GWAS on half of our AJ IBD samples (Methods)) and three sets of variants (high impact rare sites, common variants and both sets combined). As shown in Fig. 4b, high impact rare variants exhibited better predictive power compared to common variants under both GWAS statistic sets. The areas under the curve (AUC) of rare variants were 0.754 and 0.764 for Hui et al. ^45^ and our GWAS statistics respectively, whereas the AUCs of common variants were 0.744 and 0.726, respectively. The GWAS summary statistics generated by half of our datasets generally provided better AUCs compared to the other GWAS summary statistics, probably due to higher genomic coverage. As an alternative PRS method for comparison, we built a deep learning model to predict high risk individuals using the same high impact rare sites as the above model. We trained a 7-layer convolutional neural network (CNN) model (Methods), which yielded an AUC of 0.69 in 5-fold cross-validation. The power of the deep learning approach was likely restricted by the available sample size, and our sample size may be better suited for machine learning approaches such as Random Forest. The results indicate that high impact rare variants can provide predictive power which is equivalent to common variants in identifying individuals at high risk for IBD.

In conclusion, we conducted the first large-scale study of a rare and high-impact genetic architecture in AJ IBD patients. The gene burden SKAT yielded 268 significant IBD genes, that we further prioritized with pathway enrichment and biological relevance approaches, identifying 9 plausible IBD candidate genes, two of which are well known IBD genes and 7 are novel genes. We further validated these candidate genes by RNA-seq and scRNA-seq analyses. Five high impact variants within these genes were identified as significant novel IBD-causing plausible variants. We found that adult IBD under the rare and high impact genetic architecture displays similar genetic signals as VEO-IBD. PheWAS analyses on UK Biobank samples revealed potential relatedness to IBD and other complex traits. Moreover, we employed high impact rare variant derived PRS analyses to differentiate IBD cases from healthy controls, which displayed a promising power to identify individuals at risk of IBD. These findings provide new insights into the etiology of rare and high impact mutations underlying inflammatory bowel disease in the Ashkenazi Jewish population.

## METHODS

### Sample collections

The NIDDK IBD Genetics Consortium (IBDGC) recruited samples through the following research centers: Cedars Sinai Medical Center, Icahn School of Medicine at Mount Sinai, Montréal-Boston Collaborative IBD Genetic Research Center, Johns Hopkins Genetic Research Center, University of Pittsburgh and University of Toronto, and a subset of Jewish controls from Cedars-Sinai Medical Center were obtained from The National Laboratory for the Genetics of Israeli Populations at Tel-Aviv University.

Samples were collectively sequenced at the Broad Institute. We received a total of 9,076 samples across two dataset releases (3,822 and 5,254, respectively). Samples consisted of mixed populations, but the majority were broadly of European descent.

### Read mapping and genotype calling

The raw sequence reads (Fastq files) were mapped to the reference genome by the Burrows-Wheeler Alignment (BWA) tool^46^. Mapped reads were passed to GATK to mark duplicated reads and were sorted in BAM format^47^. Next, local realignment was conducted around indels to clean up ‘SNP-like’ artifacts caused by mismatching bases introduced by alignments on the edge of indels. Lastly, the base quality score quality recalibration (BQSR) was performed to recalibrate inaccurate/biased quality estimates provided by the sequencing machine. The recalibrated bam files were passed to the variant discovery process to obtain highly credible variants stored in VCF files. During this procedure, any potential variants were called by HaplotypeCaller in GATK with GVCF model and gVCFs of single samples were merged into a single gVCF to perform GATK joint calling, which generates a raw VCF including all variants and indels. Finally, the variant quality score recalibration (VQSR) was applied to raw VCF to generate a new VCF containing high-quality variants calls. VQSR has two main steps; the first uses machine-learning to assign a well calibrated probability to each variant in raw VCF. This score is then used as cutoff to extract high quality variants. This process is run in two iterations with the SNP model and indel model, respectively. The final VCF is used for downstream analyses.

### Quality control

Several quality control processes were employed to ensure high quality genotype, and samples were used in the SKAT-O analysis. Samples were excluded for the following criteria: greater than 3% missing genotypes; discordance between inferred gender based on genotype and self-reported gender; duplicated samples as identified with KING^48^; proportion of samples identical by descent > 0.185. In addition, principal components were calculated (PLINK1.9^49^) and samples were removed if they were found to be statistically lower than the specific Ashkenazi Jewish proportion (details are described in ‘PCA and STRUCTURE analyses’ section). Variants were removed on the basis of the following criteria: MAF > 1%, only rare variants were retained for high impact variants aggregating burden analysis; significant difference between missingness in cases compared with controls; genotype rate < 95% across samples, low average depth, extreme deviation from Hardy–Weinberg equilibrium (*P* < 1×10^−6^). All quality control filtering was performed using PLINK1.9^49^ and R.

### RNA-seq data analyses

Full biopsies from the terminal ileum were collected from 302 newly diagnosed individuals under the age of 17 from the RISK cohort (GEO accession GSE57945)^50^. Samples were barcoded up to 12 per lane and sequenced using the Illumina HiSeq 2000. RNA-seq reads were mapped using TopHat2^51^ (to the human reference genome version 19). Approximately 20 million reads were successfully mapped for each individual. Following RNA-seq mapping, expression levels at the gene and isoform levels was determined and expression quantified using Cufflinks^52^ to generate FPKM estimates and HTseq^53^ to generate raw read counts. We used the R package DESeq2^54^ to determine the significance of differential expression in RNA-seq samples collected from the terminal ileum biopsies of 213 CD cases, 50 UC cases, and 35 controls of European descent from the RISK cohort.

### scRNA-seq data analyses

The details of library preparation and sequencing process have been described in a previously published work^39^. In total, we analyzed 70,226 cells from paired inflamed and uninflamed ileum from 11 CD patients. We aligned to the GRCh38 reference using the Cell Ranger v.2.1.0 Single-Cell Software Suite from 10X Genomics. The unfiltered raw matrices were then imported into R Studio as a Seurat object^55^. Genes expressed in fewer than three cells in a sample were excluded, as were cells that expressed fewer than 500 genes and with a UMI count less than 500 or greater than 60k. We normalized by dividing the UMI count per gene by the total UMI count in the corresponding cell and log-transforming. The Seurat integrated model was used to generate a combined CD model with cells from both inflamed and uninflamed samples retaining their group identity. We performed unsupervised clustering and differential gene expression analyses in the Seurat R package v.3.0.1. In particular, we used shared nearest neighbor graph-based clustering, in which the graph was constructed using from 1 to 30 principal components as determined by dataset variability shown in principal component analysis (PCA); the resolution parameter to determine the resulting number of clusters was also tuned accordingly. UMAP visualizations were produced using Seurat functions in conjunction with the ggplot2. We conducted differential gene expression analysis in each cluster using *FindMarkers* function in Seurat3.0.1 package, which performs differential expression based on the non-parametric Wilcoxon rank sum test between inflamed group vs. uninflamed group. Here, we extracted the average log expression of the 9 concerned genes across 16 annotated cell clusters.

### Ashkenazi Jewish sample identification

The Jewish HapMap dataset^56^ and 112 Europeans in the HapMap^57^ dataset were used to identify 100% Ashkenazi Jewish among IBDGC samples. Jewish samples in Eastern Europe and the Middle East and Europeans were used as a reference panel to perform PCA, aiming to validate the distribution of genetically identified AJs comparing to the AJ reference panel. Population structure analyses used 36 AJ references in Jewish HapMap datasets with all IBDGC candidates. PCA and population structure analysis were based on the same set of variants filtered by the following process: merging all IBDGC samples with all reference panels by Plink, then reducing linkage disequilibrium (LD) between markers (--indep-pairwise 50 5 0.2) by removing all markers with r^2^>0.2 (window size 50, step size 5)^58^, as well as markers in known high LD regions. Variants with MAF>0.02^59^ and genotyping rate > 95% across dataset (excluding A/T, C/G mutations) and passing above conditions were employed in PCA and STRUCTURE analyses. In population structure analyses, we removed Africans and Asians from IBDGC samples. Only ‘White’ samples, which include self-reporting AJ, self-reporting mixed AJs and European were used as candidates (Supplementary Fig. 1). Accordingly, K was set to 3 to represent AJs, mixed-AJs and Europeans to run fastSTRUCTURE. The lowest AJ proportion (0.625) in the Ashkenazi reference panel was taken as AJ cutoff, and any IBDGC candidates passing this threshold were labeled as genetically identified AJs. We validated the genetically identified AJs from fine scale PCA plots without non-European populations. An independent AJ cluster can be seen from the validation PCA plots, which is overlapped with AJ reference panel (Supplementary Fig. 2). All genetically identified AJs were plotted as sky-blue points in PCA plots.

### Variant annotation

Multiallelic sites were split into single variants using bcftools before annotation. Variant Effect Predictor (VEP, v90)^60^ and SnpEff (v4.2)^61^ were employed for annotation. We used a Python script to manage a parallel running of two annotation methods and merging of results at variant level by removing redundant annotation results. CADD score (v1.3) was added into final results. All annotation processes were conducted based on GRCh37 genome coordinates.

### High impact rare variant filtration

We retained rare and high-impact genetic variants using the following criteria: (1) Maintained variants with DP>10, MQ>40 in VCF file to control the base quality. (2) Utilized Variant Effect Predictor (VEP) to determine the effect of all variants. Variants were filtered by ‘consequence’ of VEP annotations, high impact variants were retained by virtue of their impact on genome functions: ‘missense variants’, ‘start lost’, ‘stop lost’, ‘stop gained’, ‘splice_acceptor_variant’, ‘splice_donor_variant’, ‘inframe_insertion’, ‘inframe_deletion’, ‘protein_altering_variant’, ‘start_retained_variant’, ‘stop_retained_variant’ and ‘frameshift_variant’. (3) Removed variants with MAF>0.01 according to gnomAD AJ allele frequency. When gnomAD AJ allele frequency was missing for a given variant, we used its allele frequency from our AJ cohort. (4) Employed Mutation Significance Cutoff (MSC)^62^ to control the false-negative rate of predicted deleterious mutations by well-established predictors, like CADD, SIFT and Polyphen-2. Here we retained variants with a CADD^63^ score larger than the lower boundary of 95% confidence interval of the corresponding gene’s pathogenic mutation’s CADD score. (5) Genes that are highly mutated in healthy individuals are unlikely to be disease-causing. Therefore, an estimate of accumulated mutational damage of each human gene can be particularly helpful in filtering out genes that are irrelevant for disease or phenotype. Gene damage index (GDI)^64^ is an indicator to identify highly damaged genes, and is an effective tool for filtering out variants harbored in highly damaged genes that are unlikely to be disease-causing. Only variants in genes with GDI < 13.34 were retained for further study. (6) Variants frequent in a given exome cohort, but absent or rare in public databases, have also been reported and treated as non-pathogenic variants (NPV)^65^. We removed all variants that were described in the precalculated ‘blacklist’. The remaining variants were used for further analyses.

### Association analyses

We performed SKAT-O analyses on aggregations of high impact variants to test associations between genes and IBD (CD/UC) disease status. Single variant association tests were conducted by SKAT-O as well. SKATBinary_Single function was used for single variant tests for binary traits with Firth and efficient resampling. We filtered variants using gnomAD AJ MAF<0.01. However, a few sites exceeded a MAF of 0.01 among our AJ cohort, which added extra power to their corresponding genes in the SKAT association tests. Therefore, we conservatively filtered out these variants from SKAT-O analysis by adding a ‘maf < 0.01’ parameter in SKAT functions. Model based association test was conducted by Plink in validating pathway-derived high impact variants; odds ratio and *P*-values were obtained from logistic regression running in Plink1.9.

### Raremetal meta analyses

Raremetal^40^ analyses were conducted to validate the SKAT significant genes (*P* value < 0.01). Significant genes in raremetal analyses were compared with SKAT analyses to check which genes were replicated in meta analyses. Raremetalworker was used to calculate a single variants’ statistical summary for our datasets 1 and 2, respectively. As with the SKAT-O analysis, only high impact variants in each dataset were investigated. Next, we ran Raremetal to collect statistical results on independent datasets from raremetalworker with SKAT meta function. The aggregations of high impact variants were supplied in this process to perform gene level metal analyses. SKAT function was used as association test method in running Raremetal. This process was conducted on the CD and UC analyses, respectively.

### Phenome-wide association analysis

To evaluate the potential pleiotropic effects for high impact SNPs in 268 IBD associated genes from SKAT-O analysis, we performed PheWAS using genotypes and phenotype data from the UK Biobank database containing whole genome sequencing data and EHRs for all participants. We selected the phenotypes having at least 50 cases according to the ‘Diagnoses of main ICD-10’ (Data Filed 41202 in UK Biobank showcase). In 1,643 sites from 268 IBD associated genes, 964 sites have been covered by the whole exome sequencing dataset. These overlapping SNPs have been interrogated with filtered phenotypes. The PheWAS package was used to perform the analysis in R. Bonferroni adjustment was applied to the *P*-values to identify significant associations.

### Polygenic risk score and machine-learning prediction

Polygenic risk scores (PRS) generate quantitative metrics of individuals based on the cumulative effects of risk alleles. It can simply be a summation of number of risk alleles across associated genes or accumulation of risk variants weighted by size effect. Traditionally, genome-wide significant sites have been employed to generate PRS. Here, we aimed to keep all selected variants in PRS derivation. We calculated PRS for each individual based on selected high impact variants by using the LDpred algorithm^66^. Unlike variant pruning approaches, LDpred infers the posterior mean effect size of each variant by using a prior on effect sizes and LD information from an external reference panel. We used two different external panels to compare the consequence on classification models; one is Hui et al. ^45^ AJ IBD GWAS summary, the other is GWAS statistics on half of our IBD cases and controls; the remaining set of IBD cases and controls was used as a validation dataset. Variants with ambiguous strands (A/T, C/G) were removed from all high impact sites in the validation dataset. There were seven PRS generated because of the fraction *p* of non-zero effects in the prior (1, 0.3, 0.1, 0.03, 0.01, 0.003, 0.001). All seven PRS were used as features in a Random Forest classification model, the average AUC of 10-fold cross-validation was used to compare prediction performance.

We then applied a multi-layer feedforward artificial neural network, also known as convolutional neural network (CNN), to build the prediction model using all the rare high impact variants as described above. Grid search was performed to determine the best parameter settings including numbers of hidden layers, number of neurons in each layer, activation functions of the layers, dropout ratio as well as parameters for L1 and L2 regularization. 10-fold cross-validation was performed to estimate the AUC of the tuned model (7-layer CNN model with dropout ratio of 0.19, L1 of 0.002 and L2 of 0.009).

### Pathway and enrichment analyses

We used several independent pathway and enrichment methods to obtain an IBD candidate gene list using known IBD genes as a reference. The final highly credible gene list was finalized by extracting intersection genes across IBD gene sets resulting from each pathway and enrichment analyses. The candidate genes were defined as genes which pass the relaxed threshold *P* < 0.01 in a SKAT-O IBD case control study. 268 genes were obtained from IBD-specific SKAT-O case control association studies. The IBD known genes were collected from studies summarizing IBD, CD and UC genes and fine mapping efforts of identified IBD loci harboring associations mapped to single variants with greater than 95% certainty^3,67^, which forms a list of 157 IBD associated genes.

#### Ingenuity Pathway Analysis

First, we ran Ingenuity Pathway Analysis (IPA) on the IBD candidate genes, where’Cancer’,’Organismal Injury and Abnormalities’ and ‘Gastrointestinal Disease’ ranked as the top three most correlated diseases for the input genes. To select genes most relevant to IBD, we used the’Gastrointestinal Disease’ panel to extract genes belonging to its sub-phenotypes: ‘inflammation of gastrointestinal tract’, ‘inflammation of small intestine’, and ‘colitis’. We repeated the process on CD-specific and UC-specific genes.

#### ToppGene

We used ToppGene to select candidate genes from SKAT-O significant genes. ToppGene can prioritize candidate genes based on functional similarity to a training gene list. Here we used known IBD genes for training; therefore, all IBD candidate genes were ranked by training model. Each gene was assigned scores and *P* values representing functional similarities with known genes in GO terms, disease phenotypes, pathways, etc. We retained genes with *P* values < 0.05 from candidate genes as the IBD gene list.

#### GIANT

Gene function module contains clusters of genes which have similar biological functions or shorter biological distance with each other. We used GIANT function to obtain function modules from our candidate genes. GIANT applies community detection to find cohesive gene clusters from a provided gene list and a selected relevant tissue. The most IBD relevant module was selected according to their immunologic function with global tissue condition. The non-redundant genes within these identified function modules form the IBD gene list from GIANT.

#### The Human Gene Connectome (HGC)

The HGC is the set of all biologically plausible routes, distances, and degrees of separation between all pairs of human genes. A gene-specific connectome contains the set of all available human genes sorted on the basis of their predicted biological proximity to the specific gene of interest. Here, the known IBD genes are the genes of interest; for each known IBD genes, we calculated the distances to every other known IBD genes *D*_ij_, assuming that we have a SKAT-O significant gene set A *A = {a*_1_, *a*_2_, *…, a*_*m*_*}* and IBD know gene set B *B = {b*_1_, *b*_2_, *…, b*_*n*_*}*, the biological distance derived from HGC between genes from two gene sets is be represented as 

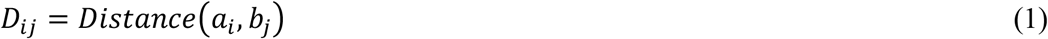

For each candidate gene *a*_*i*_ in set A, its average distance to all known IBD genes was denotes as: 

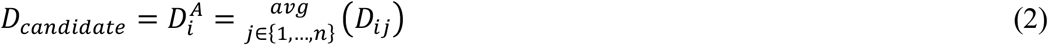

Then the overall average distance between A and B was used to represent the biological distance of candidate gene set to the known IBD gene set B: 

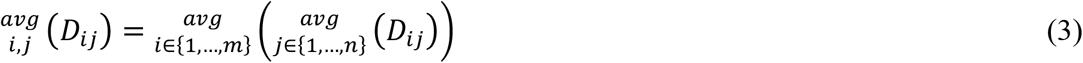

Therefore, for the known IBD gene set, we can calculate the average distance within IBD genes (*D*_*IBD*_) by checking overall average distance from gene set B to itself. For each candidate gene, if its *D*_*candidate*_ shorter than *D*_*IBD*_, it will be retained as a plausible IBD gene which contributes to the HGC IBD gene list.

At the gene set level, randomly resampling tests was conducted to demonstrate that SKAT significant genes having a shorter average biological distance to known IBD genes than random genes is not due to chance alone. For each resampling iteration, a set of genes having equal size with SKAT-O significant genes was randomly sampled from gene pool (all genes in SKAT-O inputs) and the average distances of random sets (*D*_*random*_) was calculated following equation (3). Similarly, the distance of SKAT-O significant genes was obtained (*D*_*SKAT*_) as a cutoff. The resampling tests have been conducted for 1,000, 5,000, 10,000 iterations, the *P-*value representing the number of iterations in which random sets have shorter biological distance compared to the SKAT-O significant gene set (*D*_*random*_ < *D*_*SKAT*_) among all iterations in each resampling process.

### Gene prioritization based on pathway analyses results

The SKAT-O significant IBD genes (P < 0.01) were prioritized by their biological importance in IBD pathways. One gene may be involved in multiple pathways or IBD gene function modules where other IBD genes exist as well. The biological importance was measured by counting the total number of IBD known genes in significant pathways or function modules, resulting from each enrichment analyses. The biological importance scores were added up as the final score to prioritize genes. Specifically, we collected gene sets for pathways and gene function modules from the following pathway/function analyses: (1) InnateDB^68^ pathway analysis: pathways with *P* value < 0.05. (2) InnateDB gene ontology analysis: gene list with *P* value < 0.05. (3) Networkanalyst^69^: first degree genes to each candidate genes were interrogated for counting IBD known genes. (4) IPA: canonical pathways with *P* value < 0.01 were employed. (5) Human Gene Connectome: *P* value < 0.01 for biological distance were collected in order to calculate the number of IBD genes.

## Data Availability

All data and code used in this study is available from the authors upon request.

## Acknowledgements

This research has been conducted using the UK Biobank Resource, Application 53074. We thank Gillian Belbin, Eimear E Kenny, Michael Preuss, Yashoda Sharma, Amanda Dobbyn, Ron Do, Kumardeep Chaudhary, Lisheng Zhou and Arden Moscati for research advice and administrative support. This study was funded by NIH grant R01-DK123530-01, U01-DK062413, P01-DK046763, U24-DK062429, U01-DK062422 and the Charles Bronfman Institute for Personalized Medicine, Icahn School of Medicine at Mount Sinai. We also thank for the support from Leona M. and Harry B. Helmsley Charitable Trust and Sanford Grossman Charitable Trust. D.N.C. and P.D.S. gratefully acknowledge financial support from Qiagen, Inc., through a license agreement with Cardiff University.

